# False-positive detection of Group B Streptococcus (GBS) in chromogenic media due to presence of *Enterococcus faecalis* in High Vaginal Swabs

**DOI:** 10.1101/2021.12.17.21267936

**Authors:** Abhishek Singh, Atahar Husein, Salomi Singh, Vikas Ghattargi, Dhiraj Dhotre, Yogesh S. Shouche, Stacy Colaco, Vivek Abhyankar, Suyash Patekar, Karisma Chhabria, Sushil Kumar, A.D. Urhekar, Deepak Modi

**Author notes:** **Corresponding author Dr. AD Urhekar**, Department of Microbiology, MGM Institute of Health Sciences, Kamothe Navi Mumbai, India, E mail.

## Abstract

Vaginal colonization of Group B Streptococcus (GBS) is associated with preterm births and neonatal sepsis. Thus, routine screening of GBS in prenatal care is recommended. Chromogenic media are useful in rapid and sensitive screening for GBS. herein, we evaluated the performance of Carrot broth for the detection of GBS in vaginal swabs of pregnant women. In all 20/201 (9.9%) vaginal swab samples were positive in the carrot broth. 17/20 (85%) and 19/20 (95%) samples yielded colonies on Blood agar and Crome agar respectively. However, 16s rRNA sequencing revealed that none of the carrot broth positive cultures had sequence similarities to the *Enterococcus faecalis* and not GBS. Furthermore, *Enterococcus faecalis* was detected by PCR in DNA isolated from the corresponding uncultured vaginal swabs samples, while GBS could be detected by PCR only in 4 samples. Thus carrot broth-based culture can lead to false-positive detection due to the presence of *Enterococcus faecalis*.

## INTRODUCTION

Group B Streptococcus (GBS) also known as *Streptococcus agalactiae* is a β-haemolytic, gram-positive bacterium that is considered to be one of the most important causative agents of vaginal infection and neonatal sepsis (Ellem et al., 2017, Shrestha et al., 2020). In 10 to 40% of pregnant women, there is GBS in the vagina (Shrestha et al., 2020 Ashary et al., 2020) and this has emerged as an important cause of maternal morbidity and neonatal mortality (Shabayek and Spellerberg, 2018). Every year, due to GBS infection 1 in 10 pregnancies, premature rupture of membranes (PROM), preterm births, chorioamnionitis, postpartum endometritis, urinary tract infections and post-caesarean febrile illness occurred in mother. In newborn and young infant’s sepsis, meningitis, and pneumonia is also commonly associated with maternal GBS infection (Assefa et al., 2018, Edwards et al., 2019, Tano et al., 2021). Thus, prenatal detection of GBS in vaginal swabs is recommended because GBS-positive women are eligible candidates for prophylactic intrapartum antibiotic.

GBS culture is normally performed on 5% Sheep Blood Agar (SBA) and colonies are detected by the breakdown of red blood cells that produce a characteristic zone of haemolysis. A confirmatory test for GBS is the Christie Atkins Munch Peterson (CAMP) test, or serotyping via an agglutinin reaction on selected β-haemolytic colonies (Rashwan, 2020, Xie et al., 2016). As an alternative to Blood agar, Crome agar is also used to detect GBS. However, selective enrichment of GBS in Todd-Hewitt broth before subculture on Sheep blood agar as well as Crome agar Strep B allows for efficient detection of GBS in higher vaginal swab samples (Ashary et al., 2020, Konikkara et al., 2013, Nomura et al., 2006). While enrichment of the sample and the culture are the gold standard methods for the detection of GBS in clinical samples (Lodolo et al., 2014, Rashwan, 2020), they are time-consuming and require more than 72 hours for reporting. This poses a substantial problem in the clinical setup especially in pregnant women with PROM or preterm births. This limitation has fuelled the development of liquid chromogenic media protocols, in which the swabs are directly placed in the culture medium and a colour change is indicative of a positive reaction (Konikkara et al., 2013, Trotman-Grant et al., 2012). The Carrot Broth based assays, are getting popular for sensitive and specific detection of GBS, this assay is one of the most widely used rapid culture-based screening tools for GBS in the clinical setup. Studies have demonstrated its good concordance of carrot broth assay with conventional culture-based assay systems (Berg et al., 2013, Church et al., 2008) and the prevalence of GBS using carrot broth assay is estimated to be 18% with some population differences (Church et al. 2008).

To evaluate the sensitivity and specificity of Carrot Broth in screening pregnant women in India for the presence of GBS, we initiated a prospective study in our clinic. Our results revealed that the presence of *Enterococcus faecalis* in vaginal swabs leads to false positive reporting of GBS using Carrot Broth, if additional test are not done.

## MATERIALS & METHODS

The study was approved by the Institutional Ethical Review Committee for Research on Human subjects of MGM Institute of Health Sciences kamothe Navi Mumbai (MGMIHS/RES/02/2017-18/193). We conducted this study at the Department of Microbiology & Obstetrics and Gynaecology, MGM Medical College, Kamothe, Navi Mumbai. High Vaginal swab samples were collected from pregnant women admitted to M.G.M hospital after taking the consent from the women and their families. Pregnant women consenting to participate were included in the study. Women having any antibiotic intervention less than 3 weeks prior to sample collection, and those with HIV, tuberculosis, systemic infection, were excluded from the study.

### Sample collection

Three high vaginal swabs (Hi-Media Laboratories, India) were collected from the posterior fornix using sterile swabs by an experienced gynaecologist. The first swab was placed immediately in the Carrot Broth and incubated at 37□. The second swab was placed in a lysis buffer for DNA extraction and stored at -20°C. The third swab was used for Gram stain to detect the presence of Gram-positive cocci, pus cells, and clue cells.

### Sample Processing

For GBS culture, swabs were inoculated in the Carrot broth medium (Strep B Carrot Broth™ One-Step, Hardy Diagnostics, U SA) at 37 °C for 18 to 36 hours for incubation. Positive tubes changed the colour of the broth from pale yellow to orange after 24-36 hours of incubation at 37°C while tubes that did not show colour change were further incubated at 37 °C and examined after the completion of 36 hours. Samples that changed the colour of Carrot Broth were labelled as GBS positive, while those that did not show the colour change were designated as GBS negative. All samples (those that showed as well as those that did not show colour change in Carrot Broth™ One-Step medium) were subculture on 5% Sheep Blood Agar plates (Hi-Media Laboratories, India) (referred to as Blood agar) and HiCrome™ Strep B agar plates (Hi-Media Laboratories, India) (referred to as Crome agar) at 35-37 °C in a 5% CO_2_ incubator for 18-24 hours.

### CAMP Test and BEA Test

The test using *Staphylococcus aureus* (ATCC25923) that enhances the lysis activity of red blood cells was streaked across the middle of a Blood agar plate; the suspected GBS strains were streaked perpendicular to the *S. aureus* strain (approximately 1-2 mm apart) along with control strains. A clinical isolate of GBS was used as a positive control. Streptococcus pyogenes (ATCC19615) was used as a negative control. The plates were incubated in a candle jar at 35-37 °C for 18-24 hours. An arrowhead-shaped zone of beta-haemolysis formed by the β-haemolysin of *S. aureus* at the junction of two perpendicular streak lines constituted a positive CAMP reaction (Guo et al., 2019). The colonies were also cultured on Bile Esculin Agar (BEA) (Hi-Media Laboratories, India) medium to confirm the isolates. After incubation for 18-24 hours at 37°C, the blackening of the media indicates a presence of *Enterococcus faecalis* (Manero and Blanch, 1999).

### 16S rRNA Sequencing

Single colonies of the bacteria that grew on Blood agar or Crome agar plates were cultured in liquid broth (carrot broth) overnight in shaking flask, The bacteria was collected by centrifugation. The genomic DNA of bacterial isolates was extracted by the phenol: chloroform method. Polymerase Chain Reaction (PCR) was performed to amplify 16S rRNA genes. Primers for 16S rRNA (Ghattargi et al., 2018) sequencing and the sequences are shown in Table 1. The amplified products were directly sequenced using the ABI PRISM Big Dye Terminator v3.1 Cycle Sequencing kit on a 3730xl Genetic Analyzer (Applied BioSystems). The sequence data obtained were assembled and analyzed using DNA sequence assembling software Lasergene SeqMan Pro (DNASTAR Inc.). The similarity search of newly generated 16S rRNA gene sequences was performed against the typed strains of prokaryotic species with valid published names available in the EzBioCloud’s database (Yoon et al., 2017).

**Table 1:**
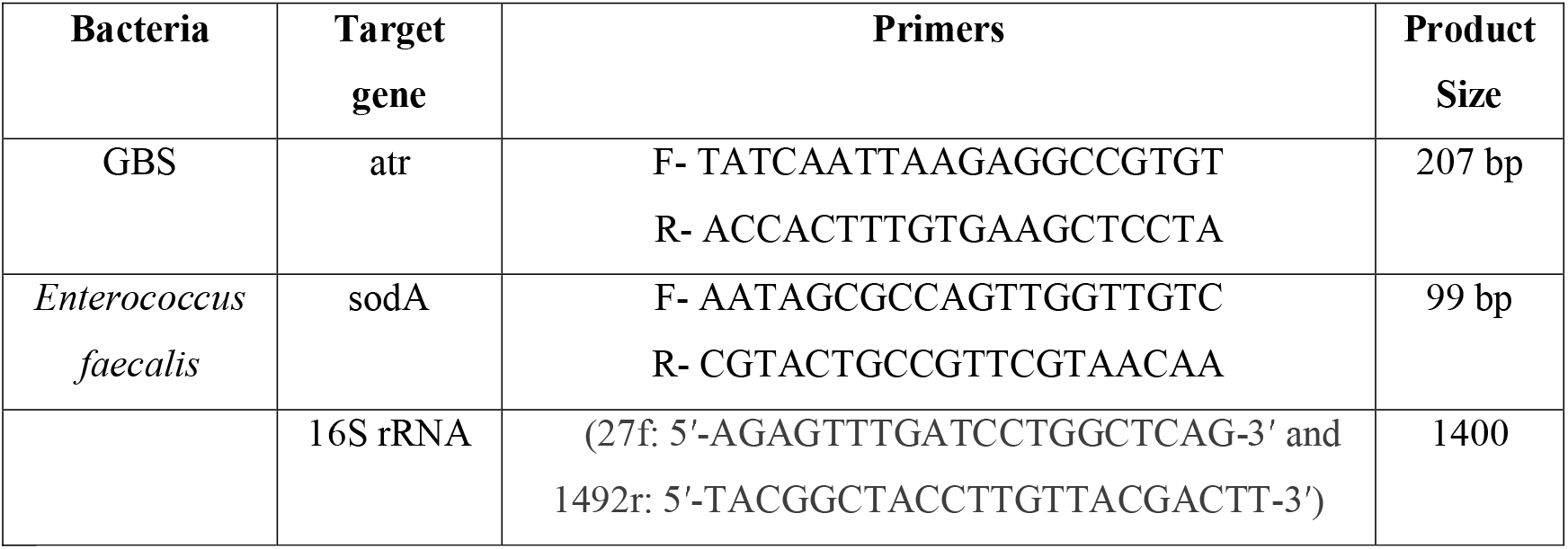
List of primers and their target genes

### Gene specific PCR for GBS and *Enterococcus faecalis*

Primers for the *atr* and *sodA* genes were designed for the GBS and *Enterococcus faecalis* respectively by using Primer3 Plus software. The specificity of primers was checked using NCBI-BLAST. (Table 1).

For gene specific PCR, individual colonies from HiCrome agar or Blood agar plates were grown in selective media and the bacterial pellet was collected. DNA was extracted from the pellet as described previously (Ghattargi et al., 2018). DNA from the swab was extracted by the spin column method (Favor Prep Soil DNA isolation Mini Kit, Favorgen, Taiwan) and stored at -20 °C. DNA extracted from each swab was analysed for the presence of GBS and Enterococcus faecalis by performing PCR for the atr and sodA genes using Taq polymerase (APSLABS, India). The PCR reaction was set as follows; initial denaturation at 95 °C for 5 minutes, denaturation at 95 °C for 30 seconds, annealing temperature at 55 °C for 30 seconds and extension at 72 °C for 30 seconds (35 cycles) in the thermal cycler (AB Applied Biosystem, Veriti). Thereafter, amplicons were run on 2% agarose gel and data were analysed accordingly.

## RESULTS

### Comparison of Carrot Broth, Blood agar, Crome agar assays for GBS Detection

Of the Table 2 compares the results of Carrot Broth positive and negative samples when inoculated on Blood agar and Crome agar cultures. 201 vaginal swabs were inoculated in carrot broth, 20 (9.9%) samples showed colour change from yellow to orange within 24 hours indicating the presence of GBS while 181 samples did not show a colour change. Of these 20 carrot Broth positive samples, 17 yielded colonies upon inoculation on blood agar and 19 yielded colonies upon inoculation in Crome agar plates. Overall, 17 of 20 carrot broth positive samples showed growth on both blood agar and Crome agar. Of the 181 Carrot Broth negative cultures, colonies were obtained in 38 samples upon inoculation on Crome agar plates. However, only 1 of the Carrot Broth negative cultures yielded colonies in the Blood agar plates; this sample was also positive in the Crome agar plate.

**Table 2:**
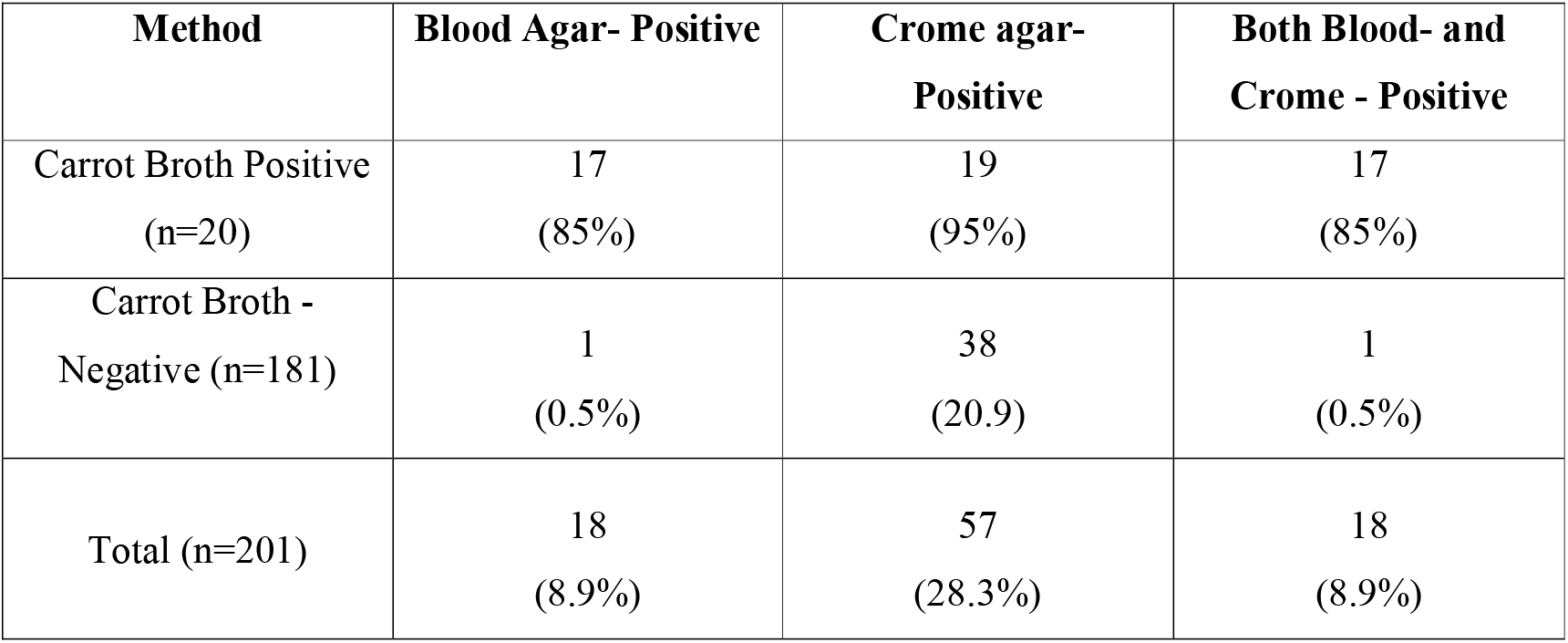
Number of high vaginal swabs of pregnant women showing growth on Carrot broth, Blood agar and Crome agar.

In all, of the 201 high vaginal swabs from pregnant women, 57 (28.3%) were positive by at least one of the three methods. Amongst these 9.9% (20/201) were positive by Carrot Broth, 8.9% (18/201) were positive upon inoculation in blood agar plates, and 28.3% (57/201) were positive when inoculated on Crome agar plates.

The CAMP test is a confirmatory biochemical assay for the identification of GBS. The standard GBS strain yielded positive results in all the assays. Of the 20 Carrot Broth positive colonies that grew on blood agar plates, 12 were positive in the CAMP assay, albeit weakly as compared to the standard strain. None of the colonies that grew on Blood agar/Crome agar plates from Carrot Broth negative cultures were CAMP test positive.

### 16S rRNA sequencing confirmed *Enterococcus faecalis* rather than GBS

For the identification of the organisms that grew in Carrot Broth, we performed 16S rRNA (V1-V2 region) sequencing of the DNA isolated from these colonies. For this, we selected 29 colonies for confirmation by 16S rRNA PCR sequencing. 20 carrot broth positive and 9 carrot broth negative samples. We found the 20 Carrot Broth positive cultures had similarities to *Enterococcus faecalis*. Of the 9 Carrot Broth negative but Crome or blood agar positive colonies, 5 were identified as *Enterococcus faecalis*, one had sequence similarity to *Escherichia coli*, two had sequences matching to *Staphylococcus haemolyticus*, and one was identical to *Acinetobacter baumannii* **(Table 3)**.

**Table 3:** Identification of bacterial species in isolates from vaginal swabs of pregnant women

| Method | **Enterococcus**<br>**faecalis** | **E.coli** | **Staphylococcus**<br>**haemolyticus** | **Acinetobacter**<br>**spp.** |
| --- | --- | --- | --- | --- |
| Carrot Broth positive<br>colonies (n=20) | 20 | 0 | 0 | 0 |
| Carrot Broth<br>negative, blood/<br>Crome agar positive<br>colonies (n=9) | 5 | 1 | 2 | 1 |

The phylogenetic tree-based comparative analysis confirmed that colonies from 25 of the 29 samples subjected to 16S rRNA sequencing had 100% similarity with *Enterococcus faecalis*. The others that had sequences divergent from *Enterococcus faecalis* were identified to be *Escherichia coli, Staphylococcus haemolyticus*, and *Acinetobacter baumannii* (Supplementary figure 1).

We also confirmed the presence of *Enterococcus faecalis* by performing gene-specific PCR. We performed semi-quantitative PCR for the specific genes (sodA) of Enterococcus faecalis with the DNA isolated from the colonies that grew on Blood agar or Crome agar plates. A single band of expected size was detected in DNA isolated from the colonies that grew on blood and crome agar plates. In contrast, no amplification was detected when we did the same with a specific gene (atr) of the GBS. The positive control had a band of the expected size and the negative control (no template) did not show amplification in any of the assays

### Biochemical Confirmation of *Enterococcus faecalis*

BEA assay differentiates GBS from Enterococci. We tested all 20 Carrot Broth positive and 18 Blood agar/Crome agar isolates by BEA along with the standard GBS strain. Amongst the 38 isolates that were negative in Carrot Broth but grew in Crome agar plates, 33 were positive in the BEA assay, suggesting their identity to be *Enterococcus faecalis* as well. Two colonies that were negative in the BEA and CAMP assays had sequence similarity to *Staphylococcus haemolyticus*. The remaining colonies (n=3) that grew on blood agar plates and were negative in the BEA assay and CAMP test had sequence similarity to *Escherichia coli* and *Acinetobacter baumannii* (1 each).

### PCR confirmed the presence of *Enterococcus faecalis* in the uncultured swab samples

To determine whether the colonies that grew on Carrot Broth/blood/ Crome agar were an artifact of a culture or indeed the primary swabs contained *Enterococcus faecalis* and/or lacked GBS, we performed PCR using primers specific for the sodA gene (99bp) of *Enterococcus faecalis* and GBS-specific (207bp) atr gene primers (Fig.1). Of the 57 vaginal swabs that were positive by culture methods, *Enterococcus faecalis* was detected in 53 swab samples by PCR. In the remaining 4 samples, 16S sequencing of the colonies demonstrated the presence of *Staphylococcus haemolyticus* (n=2) and *Escherichia coli* and *Acinetobacter baumannii* (1 each), these swabs were negative for Enterococcus.

**Fig 1:**
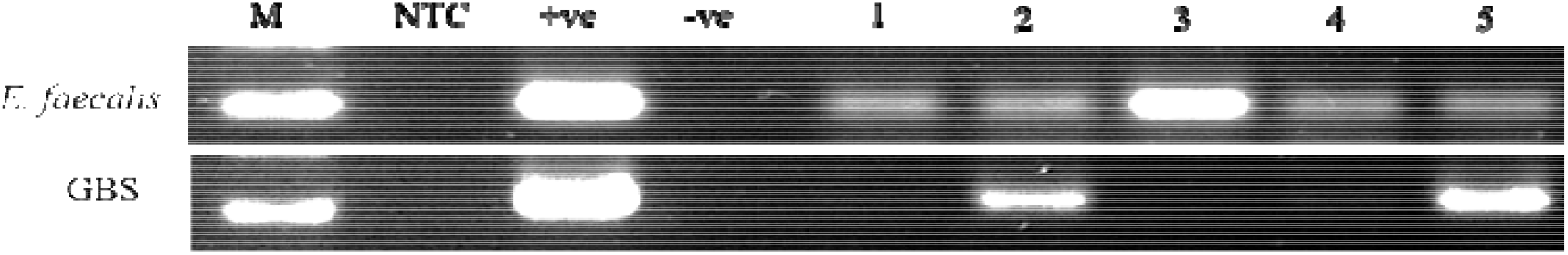
PCR for *Enterococcus faecalis* and GBS in DNA isolated from uncultured vaginal swab samples of the pregnant women. M = Maker, NTC = Non-template control, +ve control = Enterococcus & GBS standard strain, -ve control = GBS DNA for Enterococcus primer and Enterococcus DNA for GBS primer, and 1-5 = patient swab samples.

We next tested the samples for GBS by PCR. A single band of the expected size was detected in the positive control (DNA isolated from a clinical isolate). Of the 57 swabs tested for GBS by PCR, amplification was detected in 4 samples **(Table 4)**. All 4 samples were positive for GBS by PCR, indicative of co-occurrence of both bacteria. The negative controls did not show any amplicons in either case.

**Table 4:** Detection of *Enterococcus faecalis* and GBS by PCR in the uncultured vaginal swab from pregnant women

|  | **Total Positive by Culture** | **Enterococcus Positive By PCR** | **GBS By PCR** |
| --- | --- | --- | --- |
| Carrot Broth positive (n=20) | 20 | 6 (30%) | 02 (10%) |
| Crome agar (n=57) | 57 | 34 (59.6%) | 2 (3.5%) |
| Blood agar positive (n=17) | 17 | 3 (17.6%) | 0 |

## DISCUSSION

The results of the present study demonstrated that in Carrot Broth based screening tests, the presence of *Enterococcus faecalis* can lead to a false positive reporting of GBS in vaginal swabs in pregnant women, and additional tests are not used.

Vaginal colonization of GBS is associated with preterm births (Tano et al., 2021, Surve et al., 2016,) and its detection is clinically relevant for decision making towards the administration of intrapartum antibiotics (Edwards et al., 2019). According to the American College of Obstetricians and Gynaecologists, screening of GBS in the third trimester or near term is recommended in all women. Presently the gold standard method for GBS detection is a brief enrichment step (generally Todd Hewitt or Lim broth) followed by its culture in blood agar to detect haemolytic colonies (Furfaro et al., 2019). An alternate method is a culture on Crome agar post enrichment; colonies that turn blue are indicative of GBS (Manero and Blanch, 1999). The enrichment step is reported to improve the detection of GBS severalfold and reduce false negative rates (Ashary et al., 2020). While both these methods are extensively used globally, they are time-consuming, require a complete microbiological setup, and require trained personnel. Thus these methods are not readily deployable as screening tools, especially in populous countries. Thus, there are effort to develop rapid and simpler methods that do not require technical expertise. Towards this, immunologic detection of GBS antigens and liquid chromogenic media are increasingly used clinically for their ease of interpretation and fast turnaround time following sample collection (Gao et al., 2021, Guo et al., 2019).

Amongst the various choices of liquid chromogenic media, Carrot Broth is a single-step method to aid in the qualitative determination of GBS colonization in recto-vaginal swabs. This method is shown to have a sensitivity and specificity comparable or even better than the standard Lim broth enrichment followed by blood agar cultures (Block et al., 2008, Church et al., 2008). Using the Carrot Broth method, many studies have shown the prevalence of GBS in vaginal swabs in the range of 5 to 19.8% in pregnant women (Choi et al., 2021, Church et al., 2008, Xie et al., 2016). In India, the prevalence of GBS is highly variable depending on the population studied or the methods used. A recent meta-analysis carried out in pregnant Indian women has indeed shown a higher prevalence of GBS in studies using enriched media versus non-enriched media (Ashary et al., 2020). Very limited information is available regarding the performance of Carrot Broth for the detection of GBS in vaginal swabs in the Indian context (Chaudhary et al., 2017). Thus we initially aimed to determine the diagnostic performance of Carrot Broth alone and in a combination with blood and Crome agar cultures in pregnant women at term. However, during the validation, we identified that the Carrot Broth itself and in combination with blood or Crome agar cultures led to false positive results.

In our cohort, nearly 10% of women were positive in Carrot Broth, a number in range with that observed in the north Indian population reported earlier using the same method (Chaudhary et al., 2017). However, many of these positive, haemolytic colonies were either weakly positive or even negative in the CAMP test (a confirmatory test for GBS). Thus, we subjected colonies isolated from 29 randomly selected samples to 16S rRNA sequencing. To our surprise, we detected that most isolates had sequence homology to *Enterococcus faecalis* and not GBS. To determine if all the colonies were that of *Enterococcus faecalis*, we subjected them to the BEA test (Manero and Blanch, 1999). Confirming the DNA analysis results, most were BEA positive, implying that the colonies isolated from Crome or blood agar plates were that of *E. faecalis* and not GBS. To the best of our knowledge, this is the first study reporting a very high rate of false positive detection for GBS in Carrot Broth cultures due to the presence of *Enterococcus* species. Akin to our observations, there are two cases reported in the literature in which beta-haemolytic colonies obtained by the traditional method for GBS (enrichment followed by blood agar culture), when subjected to MALDI-TOF or Vitek 2 GP card, were identified as *Enterococcus* species (Savini et al., 2014, Savini et al., 2015). These reports together with our data from a large number of samples indicate that haemolysis in blood agar alone can be misinterpreted as GBS mostly due to the presence of *Enterococcus* spp.

In the identification and diagnosis of an infection, microbial culture plays a major role; however, similar kinds of biochemical reactions and cultural attributes may be conducive to more than one organism. It is known that in selective GBS broths (Todd Hewitt broth), *E. faecalis* can suppress the growth of GBS (Rosa-Fraile and Spellerberg, 2017). While similar competitive studies for Carrot Broth are not available, as *E. faecalis* is a common commensal in the vagina, we hypothesized that the co-occurrence of *E. faecalis* in high vaginal swabs may also suppress the growth of GBS, which could be the reason for the false positive detection. Towards this, we carried out PCR for both GBS and *E. faecalis* in DNA isolated from uncultured vaginal swabs from the same women that were positive by culture methods. Indeed, in four culture positive women, GBS and *E. faecalis* were present simultaneously as detected by PCR of uncultured swab DNA, 53 other uncultured swab samples were negative for GBS but were indeed positive for *E. faecalis* by PCR. Thus our results imply that the presence of *Enterococcus faecalis* in the vaginal swabs can lead to misinterpretation of the culture based tests for GBS.

In conclusion, Carrot Broth can lead to false positive reports of GBS carriage owing to the presence of *Enterococcus faecalis* in the samples. It is essential that practicing microbiologists in clinical setups are aware of this possibility and should be careful in interpreting their reports. All culture based GBS positive reports must be confirmed by not just the CAMP test but also excluded for *Enterococcus* species using the BEA test or molecular methods. More systematic studies from other parts of the world where the GBS carriage rate is high are needed to determine the usefulness of Carrot Broth in the accurate detection of GBS in a clinical setup.

## Data Availability

All data produced in the present work are contained in the manuscript

## Acknowledgment

The study was supported by MGMIHS (SEED grant). The DM lab at ICMR-NIRRH is supported by grants from Indian Council for Medical Research, The medical innovation grant (70/2/2018-MIF/BMS). KS was Nehru full-bright fellow at MCBL.

---

**Supplementary Figure 1:**
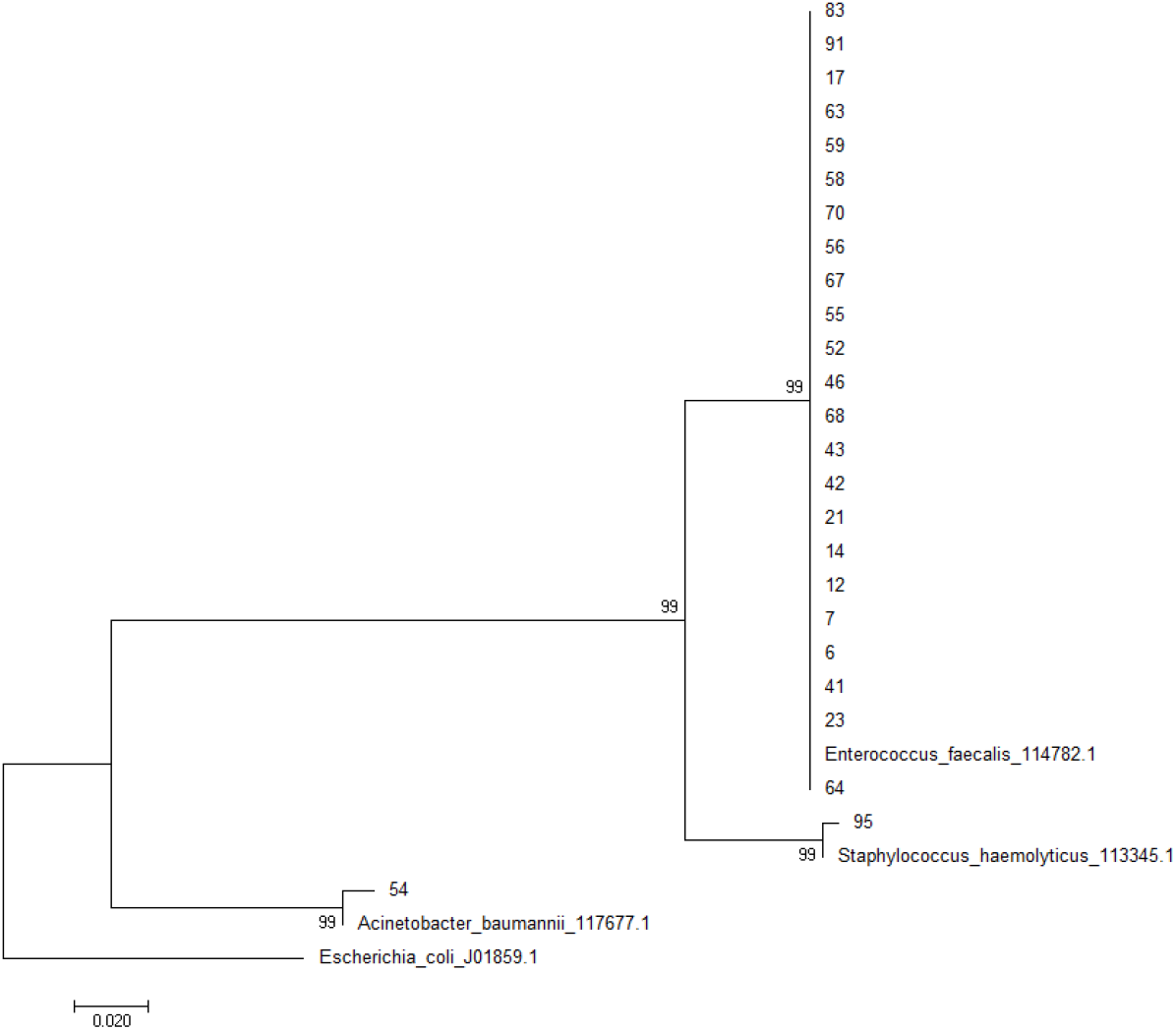
Phylogenetic tree of 16S rRNA sequences of the colonies that grew in carrot broth followed by inoculation on blood or crome agar plates. These were isolated from high vaginal swabs of pregnant women near term.

## Notes

### Competing Interest Statement

The authors have declared no competing interest.

### Funding Statement

Supported by MGM SEED Grant and ICMR Medical innovation grant

### Author Declarations

Institutional Ethical Review Committee for Research on Human subjects of MGM Institute of Health Sciences kamothe Navi Mumbai (MGMIHS/RES/02/2017-18/193), entitled "False-positive detection of Group B Streptococcus (GBS) in chromogenic media due to presence of Enterococcus faecalis in High Vaginal Swabs" approved the study.

xI have followed all appropriate research reporting guidelines and uploaded the relevant EQUATOR Network research reporting checklist(s) and other pertinent material as supplementary files, if applicable.

## References

Ashary N, Singh A, Chhabria K, Modi D. Meta-analysis on prevalence of vaginal group B streptococcus colonization and preterm births in India. J Matern Fetal Neonatal Med 2020:1–9.

Assefa S, Desta K, Lema T. Group B streptococci vaginal colonization and drug susceptibility pattern among pregnant women attending in selected public antenatal care centers in Addis Ababa, Ethiopia. BMC Pregnancy Childbirth 2018;18(1):135.

Berg BR, Houseman JL, Garrasi MA, Young CL, Newton DW. Culture-based method with performance comparable to that of PCR-based methods for detection of group B Streptococcus in screening samples from pregnant women. J Clin Microbiol 2013;51(4):1253–5.

Block T, Munson E, Culver A, Vaughan K, Hryciuk JE. Comparison of carrot broth- and selective Todd-Hewitt broth-enhanced PCR protocols for real-time detection of Streptococcus agalactiae in prenatal vaginal/anorectal specimens. J Clin Microbiol 2008;46(11):3615–20.

Chaudhary M, Rench MA, Baker CJ, Singh P, Hans C, Edwards MS. Group B Streptococcal Colonization Among Pregnant Women in Delhi, India. Pediatr Infect Dis J 2017;36(7):665–9.

Choi SJ, Kang J, Uh Y. Recent Epidemiological Changes in Group B Streptococcus Among Pregnant Korean Women. Ann Lab Med 2021;41(4):380–5.

Church DL, Baxter H, Lloyd T, Miller B, Elsayed S. Evaluation of StrepB carrot broth versus Lim broth for detection of group B Streptococcus colonization status of near-term pregnant women. J Clin Microbiol 2008;46(8):2780–2.

Edwards JM, Watson N, Focht C, Wynn C, Todd CA, Walter EB, et al. Group B Streptococcus (GBS) Colonization and Disease among Pregnant Women: A Historical Cohort Study. Infect Dis Obstet Gynecol 2019;2019:5430493.

Ellem JA, Kovacevic D, Olma T, Chen SC. Rapid detection of Group B streptococcus directly from vaginal-rectal specimens using liquid swabs and the BD Max GBS assay. Clin Microbiol Infect 2017;23(12):948–51.

Furfaro LL, Chang BJ, Payne MS. Detection of group B Streptococcus during antenatal screening in Western Australia: a comparison of culture and molecular methods. J Appl Microbiol 2019;127(2):598–604.

Gao K, Deng Q, Huang L, Chang CY, Zhong H, Xie Y, et al. Diagnostic Performance of Various Methodologies for Group B Streptococcus Screening in Pregnant Woman in China. Front Cell Infect Microbiol 2021;11:651968.

Ghattargi VC, Nimonkar YS, Burse SA, Davray D, Kumbhare SV, Shetty SA, et al. Genomic and physiological analyses of an indigenous strain, Enterococcus faecium 17OM39. Funct Integr Genomics 2018;18(4):385–99.

Guo D, Xi Y, Wang S, Wang Z. Is a positive Christie-Atkinson-Munch-Peterson (CAMP) test sensitive enough for the identification of Streptococcus agalactiae? BMC Infect Dis 2019;19(1):7.

Konikkara KP, Baliga S, Shenoy SM, Bharati B. Comparison of various culture methods for isolation of group B streptococcus from intrapartum vaginal colonization. J Lab Physicians 2013;5(1):42–5.

Le Doare K, Heath PT, Plumb J, Owen NA, Brocklehurst P, Chappell LC. Uncertainties in Screening and Prevention of Group B Streptococcus Disease. Clin Infect Dis 2019;69(4):720–5.

Lodolo L, Rossi C, Canale C, Barbaglia M, Pr G, Ghiotti P, et al. Standardization and Enrichment of Culture Medium Improve Detection of Group B Streptococci during Prepartum Screening. Journal of community medicine & health education 2014;4.

Manero A, Blanch AR. Identification of Enterococcus spp. with a biochemical key. Appl Environ Microbiol 1999;65(10):4425–30.

Nomura ML, Passini Junior R, Oliveira UM. Selective versus non-selective culture medium for group B streptococcus detection in pregnancies complicated by preterm labor or preterm-premature rupture of membranes. Braz J Infect Dis 2006;10(4):247–50.

Rao GG, Khanna P. To screen or not to screen women for Group B Streptococcus (Streptococcus agalactiae) to prevent early onset sepsis in newborns: recent advances in the unresolved debate. Ther Adv Infect Dis 2020;7:2049936120942424.

Rashwan ASSA. Assessment of different methods for diagnosis of Group B streptococci during pregnancy. Obstetrics & Gynecology International Journal 2020;11.

Rosa-Fraile M, Spellerberg B. Reliable Detection of Group B Streptococcus in the Clinical Laboratory. J Clin Microbiol 2017;55(9):2590–8.

Savini V, Franco A, Gherardi G, Marrollo R, Argentieri AV, Pimentel de Araujo F, et al. Beta-hemolytic, multi-lancefield antigen-agglutinating Enterococcus durans from a pregnant woman, mimicking Streptococcus agalactiae. J Clin Microbiol 2014;52(6):2181–2.

Savini V, Gherardi G, Marrollo R, Franco A, Pimentel De Araujo F, Dottarelli S, et al. Could beta-hemolytic, group B Enterococcus faecalis be mistaken for Streptococcus agalactiae? Diagn Microbiol Infect Dis 2015;82(1):32–3.

Schrag S, Gorwitz R, Fultz-Butts K, Schuchat A. Prevention of perinatal group B streptococcal disease. Revised guidelines from CDC. MMWR Recomm Rep 2002;51(RR-11):1–22.

Shabayek S, Spellerberg B. Group B Streptococcal Colonization, Molecular Characteristics, and Epidemiology. Front Microbiol 2018;9:437.

Shrestha K, Sah AK, Singh N, Parajuli P, Adhikari R. Molecular Characterization of Streptococcus agalactiae Isolates from Pregnant Women in Kathmandu City. J Trop Med 2020;2020:4046703.

Surve MV, Anil A, Kamath KG, Bhutda S, Sthanam LK, Pradhan A, et al. Membrane Vesicles of Group B Streptococcus Disrupt Feto-Maternal Barrier Leading to Preterm Birth. PLoS Pathog 2016;12(9):e1005816.

Tano S, Ueno T, Mayama M, Yamada T, Takeda T, Uno K, et al. Relationship between vaginal group B streptococcus colonization in the early stage of pregnancy and preterm birth: a retrospective cohort study. BMC Pregnancy Childbirth 2021;21(1):141.

Trotman-Grant A, Raney T, Dien Bard J. Evaluation of optimal storage temperature, time, and transport medium for detection of group B Streptococcus in StrepB carrot broth. J Clin Microbiol 2012;50(7):2446–9.

Xie Y, Yang J, Zhao P, Jia H, Wang Q. Occurrence and detection method evaluation of group B streptococcus from prenatal vaginal specimen in Northwest China. Diagn Pathol 2016;11:8.

Yoon SH, Ha SM, Kwon S, Lim J, Kim Y, Seo H, et al. Introducing EzBioCloud: a taxonomically united database of 16S rRNA gene sequences and whole-genome assemblies. Int J Syst Evol Microbiol 2017;67(5):1613–7.

